# Disorder- and emotional context-specific neurofunctional alterations during inhibitory control in generalized anxiety disorder and major depressive disorder

**DOI:** 10.1101/2020.10.16.20214114

**Authors:** Congcong Liu, Jing Dai, Yuanshu Chen, Ziyu Qi, Fei Xin, Qian Zhuang, Xinqi Zhou, Feng Zhou, Lizhu Luo, Yulan Huang, Jinyu Wang, Zhili Zou, Huafu Chen, Keith M Kendrick, Bo Zhou, Xiaolei Xu, Benjamin Becker

**Affiliations:** The Clinical Hospital of Chengdu Brain Science Institute, MOE Key Laboratory for NeuroInformation, High-Field Magnetic Resonance Brain Imaging Key Laboratory of Sichuan Province, University of Electronic Science and Technology of China, Chengdu, Sichuan 610054, China; Chengdu Mental Health Center, Chengdu, Sichuan, 610036, China; Department of Psychosomatic Medicine, Sichuan Academy of Medical Sciences & Sichuan Provincial People’s Hospital, Chengdu, Sichuan, 610072, China

**Author notes:** Corresponding authors Benjamin Becker, The Clinical Hospital of Chengdu Brain Science Institute, MOE Key Laboratory of Neuroinformation, No.2006, Xiyuan Ave., West Hi-Tech Zone, Chengdu, Sichuan 611731, China, Xiaolei Xu, The Clinical Hospital of Chengdu Brain Science Institute, MOE Key Laboratory of Neuroinformation, No.2006, Xiyuan Ave., West Hi-Tech Zone, Chengdu, Sichuan 611731, China, Bo Zhou, The Center of Psychosomatic Medicine, Sichuan Academy of Medical Sciences & Sichuan Provincial People’s Hospital, No.32 West Second Section First Ring Road, Chengdu, Sichuan, 610072, China. authors contributed equally (Congcong Liu, Jing Dai).

**Keywords:** Generalized anxiety disorder, Major depressive disorder, Biomarker, Emotion, Inhibitory control

## Abstract

**Background:** Major Depressive (MDD) and Generalized Anxiety Disorder (GAD) are highly debilitating and often co-morbid disorders. The disorders exhibit partly overlapping dysregulations on the behavioral and neurofunctional level, and the determination of disorder-specific alterations may promote neuro-mechanistic and diagnostic specificity.

**Methods:** In order to determine disorder-specific alterations in the domain of emotion-cognition interactions the present study examined emotional context-specific inhibitory control in treatment-naïve, first-episode MDD (n = 37) and GAD (n = 35) patients and healthy controls (n = 35) by employing a validated affective go/no-go fMRI paradigm.

**Findings:** On the behavioral level MDD but not GAD patients exhibited impaired inhibitory control irrespective of emotional context. On the neural level, no alterations were observed during the positive context, yet specifically MDD patients demonstrated attenuated recruitment of a broad bilateral network encompassing inferior/medial parietal, posterior frontal, and mid-cingulate regions during inhibitory control in the negative context. GAD patients exhibited a stronger engagement of the left dorsolateral prefrontal cortex relative to MDD patients and within the GAD group better inhibitory control in negative contexts was associated with higher recruitment of this region.

**Interpretation:** Findings from the present study suggest disorder- and emotional context-specific behavioral and neurofunctional deficits in inhibitory control in MDD in negative emotional contexts and may point to a depression-specific neuropathological and diagnostic marker. In contrast, GAD patients may maintain intact inhibitory performance via compensatory recruitment of prefrontal regulatory regions.

## Introduction

With global prevalence rates as high as 7% (1,2), depression and anxiety disorders have become one of the leading causes of disabilities (3). Comorbidity between depression and anxiety disorders is generally high, with Major Depressive Disorder (MDD) and Generalized Anxiety Disorder (GAD) exhibiting a particular high co-morbidity (4–6). On the symptomatic level both disorders are characterized by emotional and cognitive dysregulations, including exaggerated negative affect and impaired executive functions (7,8). The disorders moreover share therapeutic responsivity (9,10), genetic risk factors (5), and neural circuit disruptions (11–15), suggesting partly overlapping neurobiological pathways. On the other hand, disorder-specific phenotypes such as anhedonia (depression) and physiological hyperarousal (specific to anxiety disorders) exist (16). In line with the differential profiles on the symptomatic level, initial transdiagnostic neuroimaging studies that directly compared MDD and GAD patients revealed disorder-specific neurofunctional alterations (13–15), which are of particular importance to promote neuro-mechanistic, diagnostic and therapeutic specificity.

On the symptomatic level, key symptoms of both disorders encompass shared dysregulations in emotional and cognitive domains. Patients with MDD as well as GAD exhibit automatic, persistent and uncontrollable negative thoughts about themselves and their future (17). Neuropsychological case-control studies that compared either MDD, or GAD patients with healthy controls revealed an increased automatic attentional bias toward negative emotional stimuli (18–20), which may further exacerbate the negative emotional state (21,22). Together with the impaired capability to regulate negative emotions the exaggerated reactivity to negative emotional information and accompanying high arousal may critically impede cognitive processing in the domains of attention, memory and cognitive control.

Response inhibition represents an important core component of the cognitive control system and refers to the suppression of prepotent behavioral responses to meet current contextual and task demands (23). Previous studies revealed a lack of inhibitory control of prepotent stimulus-response contingencies across psychiatric disorders (12), suggesting a putative transdiagnostic deficit. However, emotional context- and disorder-specific dysregulations in the interplay between emotion processing and inhibitory control remain poorly understood.

Initial case-control studies examined the influence of emotional context on inhibitory control in MDD by means of affective go/no-go paradigms and reported emotional context-specific control deficits in depressive patients, such that inhibitory control deficits were predominately observed in the context of emotional stimuli. For instance, relative to healthy controls, MDD patients did not exhibit a general cognitive control deficit but presented a mood congruent bias for emotionally salient stimuli (24,25). In contrast, research on emotional context-specific inhibitory control deficits in GAD has been scarce and revealed rather inconsistent findings, with a recent case-control study reporting enhanced proactive control of negative valence distractors in GAD patients (26).

Likewise, studies employing neuroimaging methods to delineate the underlying neurofunctional basis of inhibitory control deficits in emotional contexts have mainly focused on MDD. For instance, two previous case-control neuroimaging studies employing affective go/no-go paradigms reported that MDD patients showed emotional-context specific aberrant neural engagement of frontal regions (27), and attenuated neural recruitment of the right dorsolateral prefrontal cortex (DLPFC) and bilateral occipital cortex during inhibitory control trials (no-go targets) that followed a negative, but not a positive, stimulus (28). Treatment evaluation studies in MDD furthermore reported that administration of non-invasive stimulation to frontal regions normalized emotion-specific cognitive control deficits in depression (29,30). Together, these findings suggest that aberrant emotion-cognition integration in frontal regions may underpin the emotional-context specific cognitive control deficits in MDD. In comparison, research on emotional-context dependent neurofunctional alterations in GAD is scarce. One study reported decreased right DLPFC amplitudes in GAD patients compared to healthy controls during inhibition of negative information in an explicit emotional inhibition paradigm, while the patients exhibited intact processing during an implicit emotional inhibition paradigm (31).

Overall, the above findings suggest that deficient inhibitory control in emotional contexts may represent a dysregulation that can differentiate between MDD and GAD and thus represent a behavioral and neurobiological marker with a promising potential to uncover disorder-specific pathological mechanisms. Against this background the present study employed a transdiagnostic design during which patients with MDD or GAD, and matched control subjects underwent an affective (linguistic) go/no-go paradigm with concomitant fMRI acquisition. fMRI was employed to allow the determination of the neurobiological basis of the pathology-relevant dysregulations and to additionally determine potential compensatory mechanisms on the neural level that may allow maintenance of normal task performance on the behavioral level via stronger neural engagement (13,32,33). To account for effects of treatment or progressive dysregulations during the course of the disorder treatment-naïve first episode patients were recruited. Given that recent meta-analyses reported that transdiagnostic impairments in cognitive control are neurally mediated by aberrant recruitment of the fronto-parietal cognitive control networks, as well as the anterior insula, and the midcingulate/presupplementary motor area (11,34), and that a growing number of case-control studies suggest separable and emotional context specific cognitive control alterations in GAD and MDD, we hypothesized that MDD and GAD patients manifest distinct emotional context-specific neural impairments. In particular we hypothesized that, (1) compared to controls, the MDD group would exhibit impaired inhibitory control in negative but not positive emotional contexts; and that (2) MDD patients would exhibit deficient recruitment of the frontal cognitive control network compared to controls. Given the inconsistent findings with respect to implicit emotion regulation in GAD (13,19,26) we hypothesized that GAD patients would exhibit either subtle or no alterations as compared to the healthy reference group reflecting an MDD-specific deficit in emotional-context specific inhibitory control.

## Method

### Ethics statement

The study was approved by the local ethics committee at the UESTC and adhered to the latest revision of the Declaration of Helsinki. Written informed consent and agreement to experimental procedures was obtained from all participants before enrollment.

### Participants

To control for confounding effects of pharmacological or behavioral treatment and changes related to progressive maladaptations during the course of recurrent episodes of the disorders (e.g. 35,36) treatment-naïve first-episode patients with generalized anxiety disorder (GAD, n=35) or major depressive disorder (MDD, n=37) as well as matched healthy controls (HC, n=35) were recruited. Patients were recruited at the Sichuan Provincial People’s Hospital and The Fourth People’s Hospital of Chengdu (Chengdu, China). To facilitate diagnostic accuracy a two-step approach was employed: (1) Diagnoses according to DSM-IV (Sichuan Provincial People’s Hospital) or ICD-10 (Fourth People’s Hospital of Chengdu) criteria were initially determined through clinical interviews by experienced psychiatrists, and next (2) independently confirmed by an experienced clinical psychologist by means of the Mini International Neuropsychiatric Interview (M.I.N.I.) for DSM-IV. Healthy controls (HC) were recruited via advertisements. The following exclusion criteria were applied to all participants, including HC: (1) current or history of the following axis I disorders according to DSM criteria: post-traumatic stress disorder, feeding and eating disorders, substance use disorder, bipolar disorder, and mania, (2) current or history of medical or neurological disorders, (3) acute (within six weeks before the assessments) or chronic use of medication, (4) acute suicidal ideation, (5) contraindications for MRI assessment. To facilitate data quality, and reduce the burden for the participants, participants were explicitly asked whether their current status (e.g. exhaustion, emotional state) allowed proceeding with the assessments before each assessment (e.g. MRI scanning, questionnaire assessment). The patients did not receive treatment during the period of further diagnostic clarification during which the fMRI assessments were scheduled (<5 days after admission). Demographic data, current levels of depressive and anxious symptoms were assessed by means of validated questionnaires (BDI-II, PSWQ; 37,38). Given that previous studies reported effects of childhood trauma experience on brain function and inhibitory control (e.g. 39–41) levels of traumatic experience before the age of 16 were assessed using the Childhood Trauma Questionnaire (CTQ; 42). Details are provided in **Table 1**. The study was part of a larger project that aimed at determining common and disorders-specific alterations in MDD and GAD, the affective go/no-go paradigm reported in the present study was preceded by resting state fMRI acquisition (15) and followed by a pain empathy paradigm (14).

**Table 1.** Demographics, symptom load, and early life stress

|  | GAD<br>(N=26) | MDD<br>(N=30) | HC<br>(N=34) |  |  |  |  |
| --- | --- | --- | --- | --- | --- | --- | --- |
| Male | N = 14 | N = 7 | N = 12 |  |  |  |  |
|  | Mean(SEM) | Mean(SEM) | Mean(SEM) | *F* | GAD vs HC | MDD vs HC | GAD vs MDD |
| Age (years) | 29.85<br>(1.48) | 27.97 (1.49) | 26.53 (1.59) | $F_{2,89}=1.14$ | >0.13 | >0.49 | >0.40 |
| Education (years) | 14.87 (0.69) | 13.22 (0.80) | 14.09 (0.55) | $F_{2,89}=1.34$ | >0.41 | >0.34 | >0.10 |
| PSWQ | 58.46 (2.17)<br>(N=24) | 62.34 (1.58)<br>(N=29) | 40.00 (1.59)<br>(N=34) | $F_{2,86}=50.41$<br>*** | <0.001<br>*** | <0.001<br>*** | =0.42 |
| BDI-II | 23.83 (2.09)<br>(N=24) | 32.34 (1.82)<br>(N=29) | 5.44<br>(1.00)<br>(N=34) | $F_{2,86}=80.60$<br>*** | <0.001<br>*** | <0.001<br>*** | =0.002<br>** |
| CTQ | 49.91 (2.54)<br>(N=22) | 55.29 (2.41)<br>(N=28) | 42.70 (1.67)<br>(N=33) | $F_{2,81}=9.43$<br>*** | =0.07 | <0.001<br>*** | =0.30 |

PSWQ = Penn State Worry Questionnaire; BDI-II = Beck depression Inventory II; CTQ = Childhood Trauma Questionnaire; Given that some participants did not completed all questionnaires (details see also: Demographic data and symptom load) the number of subjects that indicated the respective analysis is reported for each measure. \*\* $p < 0.01$ ; \*\*\* $p < 0.001$ , Bonferroni-corrected

### Experimental paradigm

Participants underwent a previously validated implicit affective (linguistic) go/no-go fMRI paradigm that has been developed to explore the neurocircuitry underlying the interaction between emotional context and response inhibition (43,44). The paradigm was designed as mixed event-related block design and behavioral responses were based on orthographical cues: participants were required to perform a right index-finger button press after the presentation of a word in normal font (go trial) and to inhibit this response after the presentation of a word in italicized font (no-go trial). The emotional context of response inhibition was experimentally manipulated by employing words of different valence (negative, neutral, positive) in the blocks. A total of 54 words written in Chinese were used (18 per emotional context condition). Words were matched across the valence conditions for word frequency and length (length of each word = 4 characters). A pre-study in an independent sample of n = 18 subjects demonstrated a high emotional category-specificity of the words as well as comparable imaginability, intensity (positive, negative) and word frequency between the conditions (details see **Supplementary Table S1**).

The stimuli were presented over two runs and each run comprised two blocks of the six permutations of task (go vs. no-go) and emotional context (positive, neutral, negative): neutral go (neu go), neutral no-go (neu no-go), negative go (neg go), negative no-go (neg no-go), positive go (pos go), positive no-go (pos no-go). Order of presentation was counterbalanced. Go blocks included 18 words in normal font (100% Go trials) and no-go blocks included 12 normal font words (66·7% go trials) and 6 italicized font words (33·3% no-go trials), presented in a pseudorandomized order. All blocks were preceded by a brief instruction (‘For normal font please respond by button press, otherwise, no response’) that was presented for 4s (**Fig. 1**). Each word was presented for 300ms followed by a 900ms interstimulus interval (total block duration=21·6 sec). Each block was followed by a low-level inter-block interval of 16s. Total task duration was 17min. To ensure that all participants understood the task paradigm they underwent a brief practice run with different words before the experiment.

**Fig.1.**
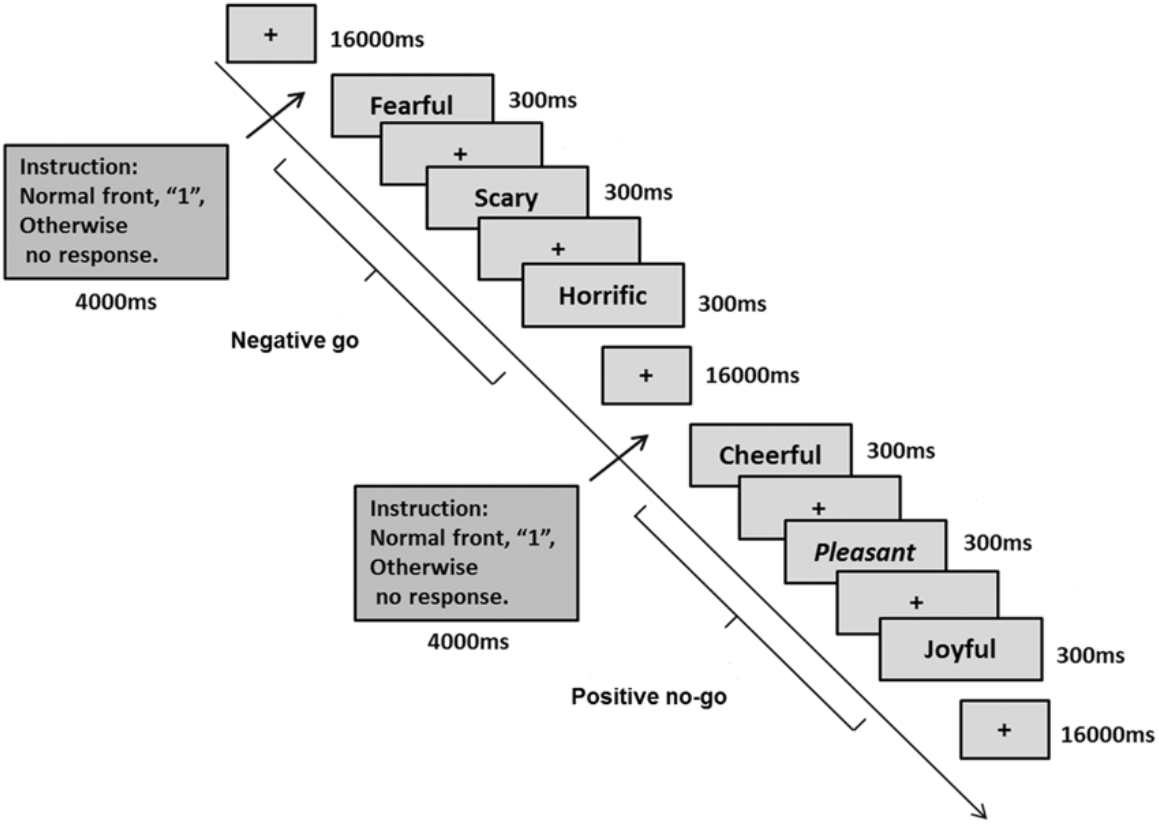
Experimental paradigm and timing: A negative go block and positive no-go block are depicted for the emotional linguistic go/no-go task.

### MRI data acquisition

MRI data were acquired on a 3 Tesla GE MR750 system (General Electric Medical System, Milwaukee, WI, USA). To exclude subjects with apparent brain pathologies and improve normalization of the functional time series, T1-weighted high-resolution anatomical images were acquired with a spoiled gradient echo pulse sequence, repetition time (TR) = 6 ms, echo time (TE) = minimum, flip angle = 9°, field of view (FOV) = 256 × 256 mm, acquisition matrix = 256 × 256, thickness = 1 mm, number of slice = 156. For the functional MRI timeseries a total of 512 functional volumes were acquired using a T2*-weighted Echo Planar Imaging (EPI) sequence (TR = 2000 ms, TE = 30 ms, FOV = 240 × 240 mm, flip angle = 90°, image matrix = 64 × 64, thickness/gap = 3·4/0·6mm, 39 axial slices with an interleaved ascending order).

### MRI data processing

MRI data was processed and analyzed using Statistical Parametric Mapping (SPM12; Wellcome Department of Cognitive Neurology, Institute of Neurology, London, United Kingdom). For each subject and run, the first 6 functional volumes were discarded to allow magnet-steady images. The remaining functional images were preprocessed using standard preprocessing procedures including: slice timing, realigning to correct for head motion, a two-step normalization to Montreal Neurological Institute (MNI) standard space including co-registration to the T1-weighted structural images and application of the segmentation parameters obtained from segmenting the structural images to the functional time-series (interpolated at 3×3×3mm voxel size) and spatial smoothing using an 8mm full-width at half-maximum (FWHM) Gaussian kernel.

For statistical analysis a two-step general linear model (GLM) approach was employed. At the single subject level an event-related general linear model (GLM) was employed including condition-specific regressors modelling the six experimental conditions and the six head motion parameters. Regressors for the experimental conditions were convolved with the default SPM hemodynamic response function (HRF). The design matrices additionally included a high pass filter to control for low frequency components and a first-order autoregressive model (AR[1]) to account for autocorrelation in the time-series. To explore distinct dysregulations in the interaction between the emotional and inhibitory control between the diagnostic categories the primary contrasts of interest [(a) for negative valence: [(neg vs. neu) × (no-go vs. go)], and (b) for positive valence: [(pos vs. neu) × (no-go vs. go)] were subjected to voxel-wise whole-brain group level ANOVA analyses followed by post-hoc voxel-wise interdependent t-tests to further determine disorder-specific alterations. In addition, to further disentangle the interaction effect and to determine inhibition-related neurofunctional alterations an additional voxel-wise ANOVA and corresponding post-hoc tests examined inhibition specific neurofunctional alterations (e.g., [neg no-go vs. neu no-go].

### Statistical analysis and thresholding

Demographic and clinical data between the groups were examined via one-way analysis of variance (ANOVA) and chi-square tests. Two-way mixed ANOVAs with emotional valence × group were conducted to analyze response times of correct Go trials. Accuracy rates were investigated using a three-way mixed ANOVA with trial category (go trials vs no-go trials) × emotional valence × group as factors. All analyses were conducted using the Statistical Package for Social Sciences, version 21 (SPSS Inc., USA). Statistical analyses were adjusted for variance non-sphericity using the Greenhouse–Geisser correction. All post-hoc analyses employed appropriate Bonferroni adjustment.

For the analyses of the fMRI data the primary contrasts of interest were subjected to group-level random-effects analysis. A whole-brain voxel-wise analysis examined differences between the diagnostic categories (MDD, GAD, and HC) using a one-way-ANOVA design (columns in the design matrix representing the GAD, MDD, and HC group) with gender and age as covariates in SPM12. Significant main effects of group were followed up by voxel-wise post-hoc independent *t*-tests that directly compared the three diagnostic groups. The voxel-wise statistics were performed on the whole brain level using a cluster-level Family-Wise Error (FWE) correction at *p* < 0·05. In line with recommendations for the application of cluster-level correction approaches an initial cluster forming threshold of *p* < 0·001 was employed (45,46).

In line with our previous studies (14,15) the categorical analysis was flanked by a subsequent follow-up dimensional analytic approach that examined associations between the observed categorical differences on the group level and MDD (BDI II scores) or GAD (PSWQ scores) symptom load, respectively, in the entire sample.

## Results

### Demographic data and dimensional symptom load

After initial quality assessment of the data 26 patients with GAD, 30 patients with MDD and 34 HCs were included in the final analysis (detailed exclusion procedure see **Supplementary figure S1**). Participants in the GAD, MDD, and HC groups were of comparable age (*p*=0·33), gender distribution (χ^2^=0·06), and education level (*p*=0·27). Some patients (one HC) reported being too exhausted to continue with the self-report questionnaire following the MRI assessments. The number of subjects for the GAD and MDD group therefore varies from 26 to 24 and 30 to 29 (BDI II, PSWQ), 26 to 22 and 30 to 28 (CTQ) respectively. Importantly, testing differences in the ratio of participants that discontinued the self-reported questionnaires did not reveal significant differences between the patient groups (Chi-square test, all *p*s>0·05, detailed numbers provided in **Table 1**). One-way ANOVA analysis for depressive symptom load revealed a significant main effect of group (BDI-II, *F*_2,86_ =80·60, *p*< 0.001, *η*^*2*^_*p*_*=* 0·66) with post-hoc analyses indicating that depressive symptom load was higher in both, GAD and MDD patients compared to HC, and in MDD compared to GAD patients (*p* values, GAD vs HC <0·001, MDD vs HC <0·001, GAD vs MDD = 0·002). Examining GAD symptom load revealed a significant main effect of group (PSWQ, *F*_2,86_= 50·41, *p*<0.001, *η*^*2*^_*p*_*=* 0·55) with GAD symptom load being significantly higher in both patient groups relative to HC, but not significantly different between the two patient groups (*p* value, GAD vs HC <0·001, MDD vs HC <0·001, GAD vs MDD = 0·62, details see **Table 1**).

### Behavioral results

Examination of response accuracy revealed a main effect of trial category suggesting that all participants responded more accurately for go trials (97·0%, *SD* = 0·6%) as compared to no-go trials (69·9%, *SD* = 1·9%; *F*_1,88_ =200·42, *p*< 0·001, *η*^*2*^_*p*_*=* 0·70). A significant main effect of emotional valance reflected that all participants made less accurate responses to positive (82·3%, *SD* = 1·1%) as compared to negative trials (84·2%, *SD* = 1·1%; *F*_*2,88*_ =4·74, *p*< 0·05, *η*^*2*^_*p*_*=* 0·10). There was a significant interaction effect between emotional valance and trial category (*F*_2,88_=5·53, *p*< 0·01, *η*^*2*^_*p*_*=* 0·11), with post-hoc tests demonstrating that all participants made less accurate responses in the pos no-go trials (67·6%, *SD* = 2·1%) as compared to the neg no-go trials (71·6%, *SD* = 2·0%, *p* = 0·005). Furthermore, there was a significant main effect of group (*F*_2,88_=5·15, p< 0·01, *η*^2^_p_= 0·11) with post-hoc tests demonstrating that MDD patients made significantly less accurate responses (78·8%, SD = 1·7%) compared to both, GAD patients (86·7%, *SD* = 1·8%, *p* = 0·007) and HC (84·9%, *SD* = 1·6%, *p* = 0·03), whereas GAD did not differ from HC (84·9%, *SD* = 1·7%, *p* = 0·69. **Fig. 2**). The main effect of group remained robust after including gender and age as covariates (F_2,87_=6·08, *p*=0·003, *η*^2^_p_= 0·13). No other main or interaction effects with respect to accuracy reached significance (*p*s > 0·16). No significant main or interaction effects were observed in the analysis of response times.

**Fig.2.**
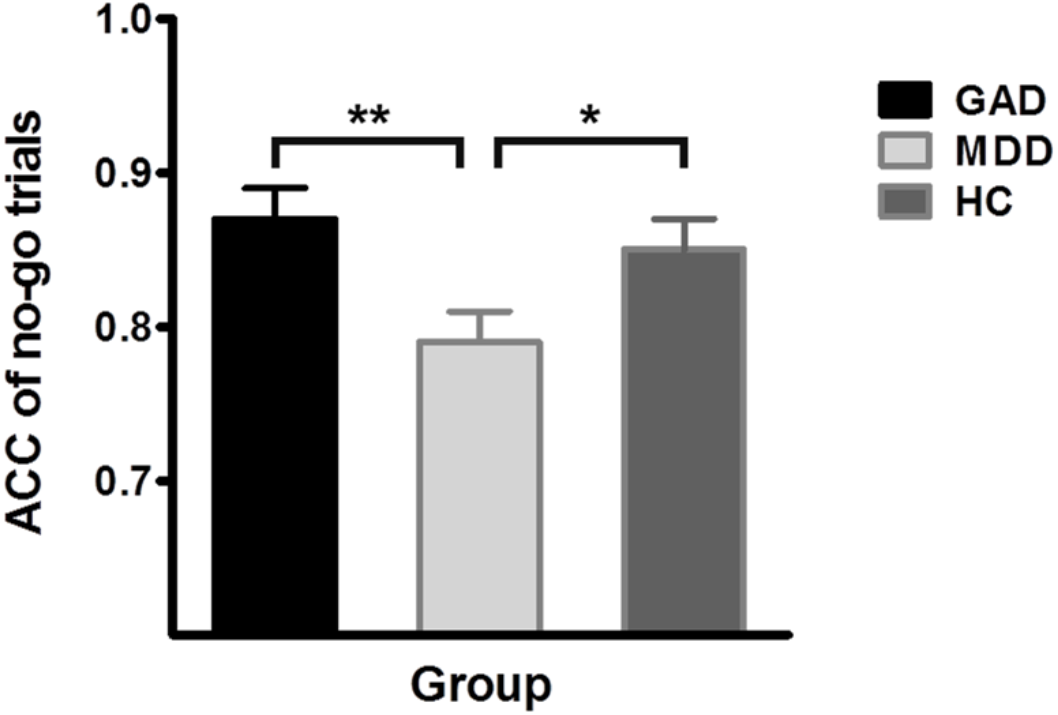
Accuracy of no-go trial in GAD, MDD and HC groups. ACC, accuracy. Error bars indicate standard error. \**p* < 0·05; \*\**p* < 0·01, Bonferroni-corrected.

### Neuroimaging results

ANOVA models were employed to determine differences between the diagnostic groups in the primary contrasts relevant to identifying the influence of emotional context on inhibitory control, specifically (a) positive context: [(pos vs. neu) × (no-go vs. go)], and (b) negative context: [(neg vs. neu) × (no-go vs. go)]. Examining the positive emotional context revealed no significant differences between the groups. In contrast, the voxel-wise whole-brain ANOVA examining the negative context revealed a significant interaction effect involving the factor group in a widespread bilateral sensory-motor and cognitive control network, encompassing the left postcentral gyrus/supramarginal gyrus (MNI [-57 -33 18], *p*_FWE-cluster_ = 0·007, k = 164, *F*_2,85_ = 13·14), right postcentral gyrus/precentral gyrus/supramarginal gyrus (MNI [69 -24 12], *p*_FWE-cluster_ < 0·001, k = 248, *F*_2,85_ = 15·03), and the bilateral middle cingulate (MNI [6 -6 51], *p*_FWE-cluster_ = 0·005, k = 176, *F*_2,85_ = 14·23; **Fig. 3**). A subsequent direct comparison of the three groups by means of voxel-wise SPM12 independent t-tests revealed significantly attenuated engagement of these regions in MDD patients compared to both, HC and GAD (details see **Fig. 4a, Fig. 4b**), whereas GAD patients exhibited no differences compared to HC, indicating that the interaction effect was driven by altered neural activation in the MDD patients. Moreover, the direct comparison between GAD and MDD groups additionally revealed significantly increased recruitment of the left dorsal lateral prefrontal cortex (DLPFC, MNI [-39 54 21], *p*_FWE-cluster_ = 0·004, k = 251, *t*_85_ = 4·34) in the GAD relative to MDD patients **Fig. 4b**).

**Fig.3.**
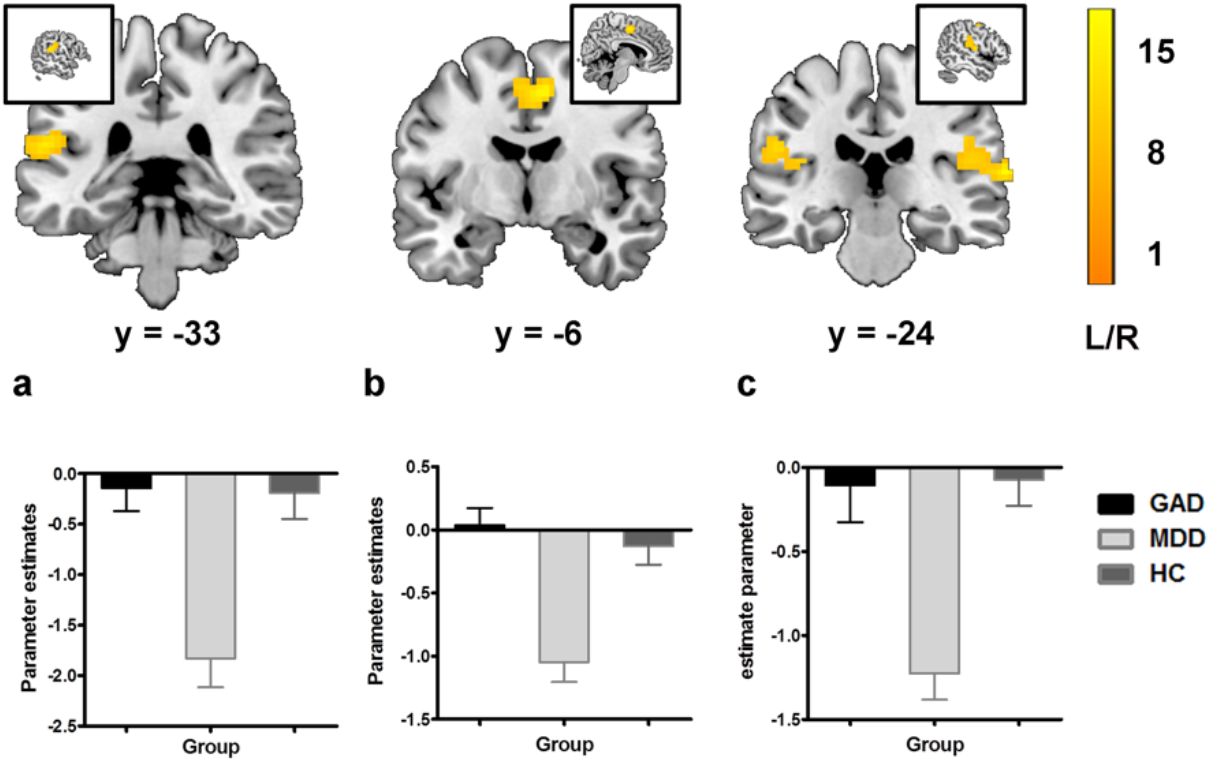
Main effect of diagnostic group (GAD, MDD and HC) for the interaction effect between negative emotional and inhibitory response [(neg – neu) × (no-go – go)]. All effects survived the family-wise error (FWE) correction for multiple comparisons (p < 0·001). The color bar codes the *F* value. For visualization, the extracted estimates for interaction effect between negative emotional and inhibitory response [(neg – neu) × (no-go – go)] are displayed for each group, left postcentral gyrus/ supramarginal gyrus (2a), right postcentral gyrus/precentral gyrus/supramarginal gyrus (2c), and bilateral middle cingulate (2b). L/R, left/right.

**Fig.4.**
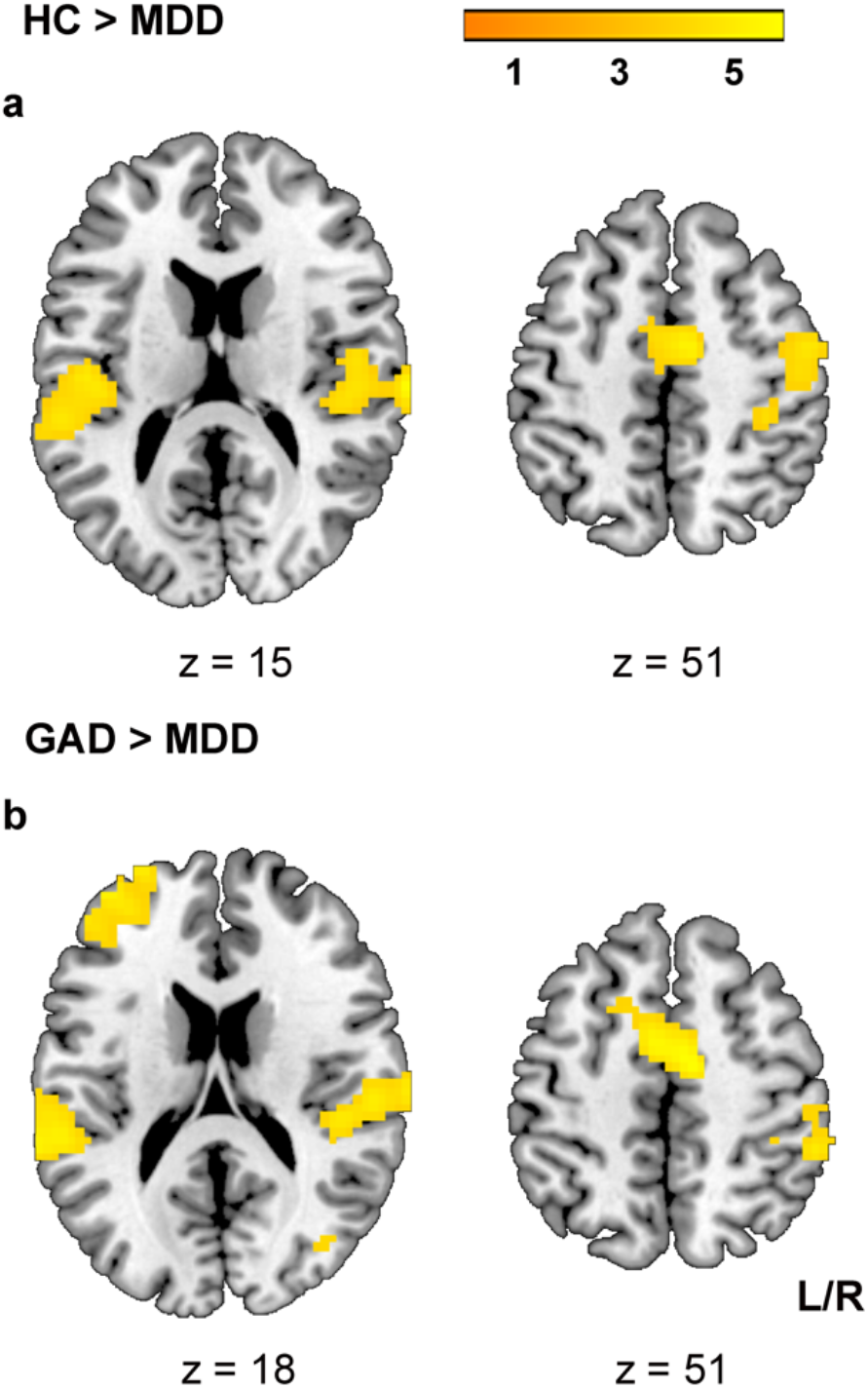
Comparisons between patients with MDD and patients with GAD and HC for the interaction effect between negative emotional and inhibitory response [(neg – neu) × (no-go – go)]. All effects survived the family-wise error (FWE) correction for multiple comparisons (*p* < 0·001). The color bar codes the *t* value. L/R, left/right.

To further disentangle the complex interaction effect and to determine alterations during inhibitory control in negative contexts an additional voxel-wise ANOVA focused on the corresponding contrast [neg no-go vs. neu no-go]. The one-way-ANOVA model in SPM12 (columns in the design matrix representing the GAD, MDD, and HC group) with gender and age as covariates revealed a significant main effect of group during inhibitory control in negative contexts encompassing the network described above (*p*s_FWE-cluster_ < 0·05, details see **Fig. 5a**). A subsequent direct comparison of the three groups by means of voxel-wise SPM12 independent t-tests revealed significantly attenuated engagement of these regions in MDD patients compared to both, HC and GAD (details see **Fig. 5b, Fig. 5c**), whereas GAD patients exhibited no differences compared to HC, further emphasizing inhibitory control-specific neurofunctional alterations in MDD.

**Fig.5.**
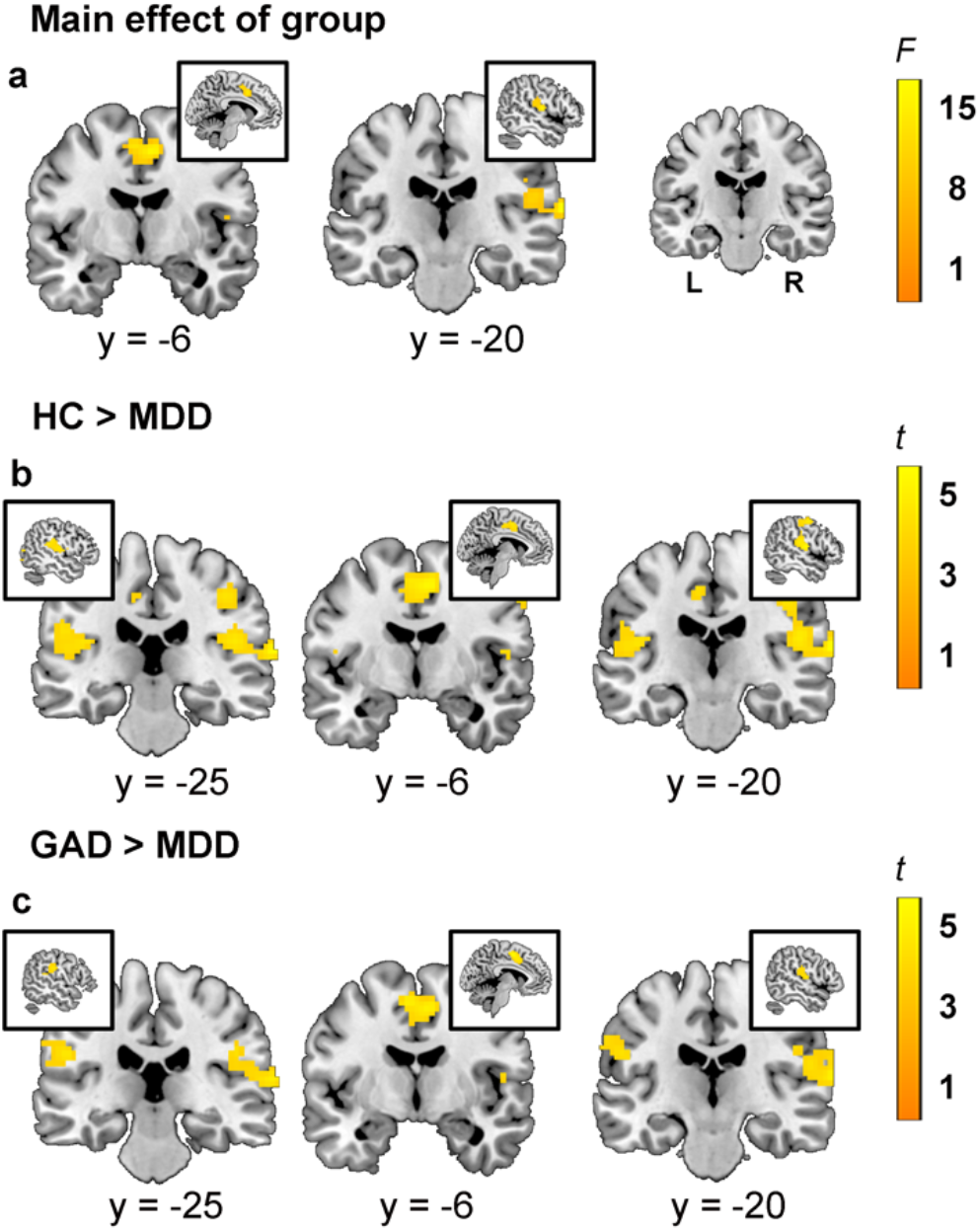
Main effect of diagnostic group (GAD, MDD and HC) and between-group results for the response inhibition within negative vs. neutral valence. All effects survived the family-wise error (FWE) correction for multiple comparisons (*p* < 0·001). L, left; R, right.

### Dimensional analysis

In line with our previous studies (e.g. 14) the identified categorical between-group differences were followed up by a dimensional analysis approach. To this end, associations between BDI II scores and the identified behavioral (accuracy), and neural alterations (extracted parameter estimates) in the entire sample, were examined by linear models (FSL PALM-alpha toolbox (https://fsl.fmrib.ox.ac.uk/fsl/fslwiki/PALM, Permutation Analysis of Linear Models, number of permutations = 10,000) including GAD symptom load as covariate. No significant associations were observed (all *p* > 0·05).

### Exploratory analysis – compensatory processes in GAD

The behavioral analyses revealed that GAD patients exhibited preserved inhibitory control in the context of negative emotion and increased DLPFC recruitment in a direct comparison with MDD patients. Together this suggests a neural compensatory mechanism which may facilitate intact response inhibition in GAD patients. To explore this hypothesis, we employed voxel-wise regression models examining associations between accuracy rates for neg no-go trials with whole-brain activation in the corresponding contrast within the GAD group (FWE-cluster level correction at *p* < 0·05, initial cluster forming threshold *p* < 0·001). Results revealed that within the GAD patients better inhibitory control in negative contexts (higher accuracy rates in the negative context) was associated with increasing activation in the left DLPFC (MNI [-48 27 24], *p*_FWE-cluster_ < 0·05, k = 104, *t*_22_ = 5·58. **Fig. 6**). On the other hand, no significant correlations were observed in the MDD and HC groups. These findings may reflect that GAD patients can maintain inhibitory control in negative contexts via compensatory recruitment of the DLPFC.

**Fig.6.**
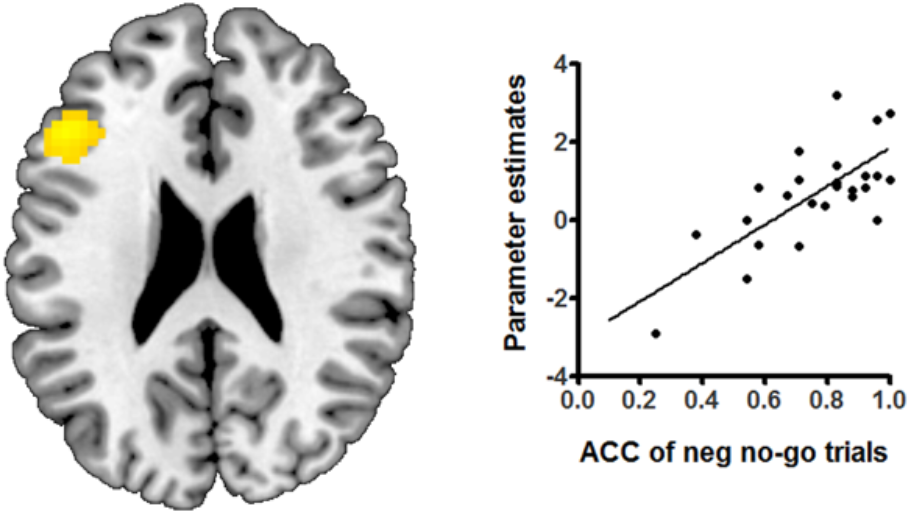
Brain activation is displayed with family-wise error (FWE) correction for multiple comparisons (*p* < 0·001). Positive correlations in the left dorsolateral prefrontal cortex (DLPFC) suggest better inhibitory control (higher accuracy rates of neg no-go trials) was associated with progressively more activation in the left DLPFC. Scatter plot on the right, associations between extracted parameter estimates and accuracy of negative no-go trials for left DLPFC. ACC, accuracy.

## Discussion

The present study aimed at determining disorder-specific behavioral and neurofunctional dysregulations in emotional context-specific inhibitory control in MDD and GAD patients. To this end we employed a validated implicit affective go/no-go fMRI paradigm in unmedicated first episode MDD and GAD patients and HC. On both behavioral and neural levels we found supporting evidence for disorder-specific impairments, such that MDD patients exhibited generally impaired inhibitory control in terms of reduced accuracy rates during no-go trials as compared to both HC and GAD patients, while GAD patients did not differ from HC. On the neural level specifically MDD patients demonstrated attenuated recruitment of a broad bilateral network encompassing inferior/medial parietal and posterior frontal as well as mid-cingulate regions during inhibitory control in the negative context, suggesting disorder- and emotional context-specific neurofunctional deficits. Further examination of disorder-specific alterations and potential compensatory recruitment on the neural level revealed that GAD patients exhibited a stronger engagement of the left DLPFC relative to MDD patients and that within the group of GAD patients better inhibitory control in negative contexts associated with increasing recruitment of this region, possibly reflecting a compensatory mechanism on the neural level that facilitates intact performance in GAD.

On the behavioral level, the observed pattern in the present study partly resembles findings in previous studies (24,25,31) such that MDD patients showed lower accuracy while GAD patients exhibited comparable accuracy with HC. However, while the previous studies reported emotion-specific inhibitory control deficits in MDD patients the present study found a general impairment in no-go accuracy irrespective of emotional context. The differences between the studies may be explained in the sample characteristics, in that previous studies examined alterations in the context of ecological and clinical validity by including MDD patients with a history of previous episodes and current pharmacological treatment (24,25) whereas the present study aimed at specifically determining disorder-specific neurobiological mechanisms while controlling for these factors. Together, the findings may indicate comparably subtle and rather general cognitive impairments during early and unmedicated stages of MDD, while emotional context-specific impairments may become increasingly pronounced during the progression of the disorder or prolonged pharmacological treatment.

On the neural level no between-group differences were observed during the positive context, yet MDD patients differed from both HC and GAD patients with respect to the recruitment of a widespread bilateral network encompassing posterior frontal, parietal, and mid-cingulate regions in the negative context. Post-hoc analyses revealed that MDD patients specifically exhibited decreased recruitment of this network during trials that required inhibitory control of the prepotent motor response, indicating specific neurofunctional deficits during cognitive control. Together with the prefrontal systems, the parietal and posterior frontal regions constitute the fronto-parietal network which has been consistently involved in cognitive control processes, including inhibition of prepotent motor responses during go/no-go paradigms (47,48). Within this network the precentral / postcentral gyrus has been specifically associated with mild emotional interference during cognitive control (49) and the parietal cortex is involved in biases relevant to stimulus-response associations, while prefrontal regions constitute a more domain-general network regulating emotional and cognitive interference (50,51).On the one hand the present findings of deficient neural engagement during cognitive control in negative contexts generally align with previous studies reporting context-specific neural alterations in MDD patients as compared to controls (28). On the other hand, these previous studies reported alterations in prefrontal regions, specifically dorsolateral and ventrolateral regions, whereas the present study found alterations in parietal, posterior frontal, and cortical midline regions, specifically the supramarginal, postcentral and precentral gyrus, as well as the MCC. The diverging results with respect to the specific location of the emotional context-specific neurofunctional alterations in MDD may be partly explained by the differences in the sample characteristics. Thus, previous studies were conducted in MDD patients with recurrent episodes of depression and under anti-depressive medication while the present study examined unmedicated first episode patients. In addition to progressive emotional and cognitive dysregulations during the course of the disorder, progressive changes in structural integrity, particularly in prefrontal regions, have been reported in MDD (52). Together with the present results this may suggest that during the progressive course of the disorder or pharmacological treatment neural alterations shift from posterior to more prefrontal regions.

MDD patients in the present study additionally exhibited deficient recruitment of the bilateral MCC during inhibitory control in the negative context. A recent transdiagnostic neuroimaging meta-analysis reported altered activity in this region as well as core regions of the fronto-parietal cognitive control network across different cognitive control paradigms and psychiatric disorders (11). A critical role of the MCC as core node for the integration of emotional context and cognitive control has been further documented in a recent study evaluating the effects of cingulotomy in patients with treatment resistant depression. The study reported that patients with treatment resistant depression who underwent focal bilateral anterior cingulotomy targeting the MCC subsequently exhibit specific impairments in recognizing negative stimuli and in inhibitory control of prepotent stimulus-response contingencies while exhibiting enhanced interference sensitivity (53). This finding suggests a critical role of the MCC for engaging cognitive control processes in the presence of negative stimuli to optimize goal directed behavior. Furthermore, recent overarching reviews suggest that – together with the anterior portion of the cingulate – the MCC constitutes a highly integrative hub bridging negative emotion processing, pain, and cognitive control with motor systems executing goal-directed behavior (53–55). Together, these findings suggest a deficient recruitment of a network engaged in the integration of emotional inference and motor systems during inhibitory control in MDD.

In contrast to previous studies that reported dimensional associations between depressive symptom-load and altered intrinsic brain architecture (15,56) as well as altered pain empathic insula reactivity (14) across GAD and MDD patients the present study did not reveal significant associations with respect to the behavioral and neurofunctional alterations observed in the categorical analysis. Together with a visual inspection of the extracted parameter estimates from the categorical approach (see **Figure 3**, lower panel), this suggests rather categorical differences between MDD and GAD in the domain of inhibitory deficits in negative contexts which may indicate a particular diagnostic specificity of dysfunctions in this domain.

A direct comparison of the patient groups with respect to neural activation during cognitive control in negative contexts further revealed higher DLPFC activation in GAD relative to MDD patients. Moreover, results from a brain-behavior analysis further revealed that within the GAD patients inhibitory control performance in negative contexts associates with stronger recruitment of this region. The DLPFC represents a core region of the domain general cognitive control network and subserves inhibitory control, cognitive flexibility, and working memory (48). Previous studies that targeted the left DLPFC with non-invasive brain stimulation techniques reported improved cognitive control in healthy individuals (57) and improved cognitive control in emotional contexts in MDD patients (29,30), suggesting that stronger engagement of this region may facilitate cognitive control performance. A pattern of intact executive performance in the context of higher neural activation following pharmacologically-induced suppression of noradrenergic signaling (32), early-onset drug use (33), and patients with obsessive compulsive disorder (58) has been previously interpreted as compensatory neural recruitment which facilitates intact performance on the behavioral level. Within this context the pattern of stronger engagement of the DLPFC in GAD patients may reflect a compensatory mechanism on the neural level that allows GAD patients at early stages of the disorder to maintain intact inhibitory performance in negative emotional contexts and furthermore suggests that fMRI may provide additional important information in diagnostic contexts by uncovering neurofunctional compensation.

Findings need to be considered in the context of limitations of the present study. Firstly, to control for important confounders such as treatment or progressive dysregulations during the course of the disorder, we employed strict enrollment criteria, which came at the cost of only a minority of patients in two large psychiatric hospitals being eligible for enrollment thus leading to a moderate sample size. Secondly, although it is suggested that there are significant differences in depression and anxiety between males and females (59,60), the relatively small sample size did not allow us to further explore gender differences. Therefore, potential gender differences need to be explored in future studies.

## Conclusion

Together, findings from the present study suggest disorder-specific neurofunctional alterations during inhibitory control in negative emotional contexts in MDD, specifically a deficient engagement of a broad bilateral network encompassing inferior/medial parietal and posterior frontal as well as mid-cingulate regions. Although GAD patients did not demonstrate deficits on the behavioral and neural level in comparison to healthy controls, stronger recruitment of the DLPFC as compared to MDD patients and associations between better inhibitory performance and higher activation of this region within the GAD group may point to a compensatory mechanism on the neural level that facilitates intact inhibitory control in GAD.

## Data Availability

The data that support the findings of present study are available from the corresponding author upon reasonable request. The data sharing adopted by the authors comply with the requirements of the funding institute and with institutional ethics approval.

## Author contributions

BB and KK designed the study and supervised the conduct of the study. XX and LL prepared the study protocols, BZ, JD, ZZ, YH, and JW performed the clinical assessments. CL, XX, YC, and FX, contributed to the data collection. BB, QZ, XZ, and HC provided methodological advice. CL, and BB performed the data analysis and results interpretation. CL, and BB drafted the manuscript, which all authors reviewed and approved for publication.

## Acknowledgments

The authors would like to express their sincere appreciation to their laboratory members and colleagues for providing valuable help. We also thank all the participants in this study.

