## supplementary information for "Disorder- and emotional context-specific neurofunctional alterations during inhibitory control in generalized anxiety disorder and major depressive disorder"

### Supplementary File

**Table S1. Results from emotional valence** (yes and no rating of the fit to the corresponding category), intensity (1-9 scale), and imaginability (1-9 scale) ratings of the word stimuli in an independent sample (n = 18) as well as word frequency.

| | Stimuli | | | | |
|---|---|---|---|---|---|
| **Measurements** | **Positive** | **Negative** | **Neutral** | ***F*** | ***p*** |
| **Emotional valence rating** | 94·58% ± 1·07% | 96·00% ± 1·07% | 96·21% ± 0·73% | 0·95 | 0·35 |
| **Frequency** | 1965·58 ± 340·04 | 1007·24 ± 446·22 | 1725·54 ± 288·05 | 1·87 | 0·16 |
| **Intensity** | 5·73 ± 0·17 | 6·15 ± 0·23 | | 1·47 (*t*) | 0·15 |
| **Imagination** | 5·76 ± 0·23 | 6·07 ± 0·22 | 5·70 ± 0·12 | 1.02 | 0.37 |

**Figure S1 Flow diagram displaying exclusion of participant and rationale for exclusion**

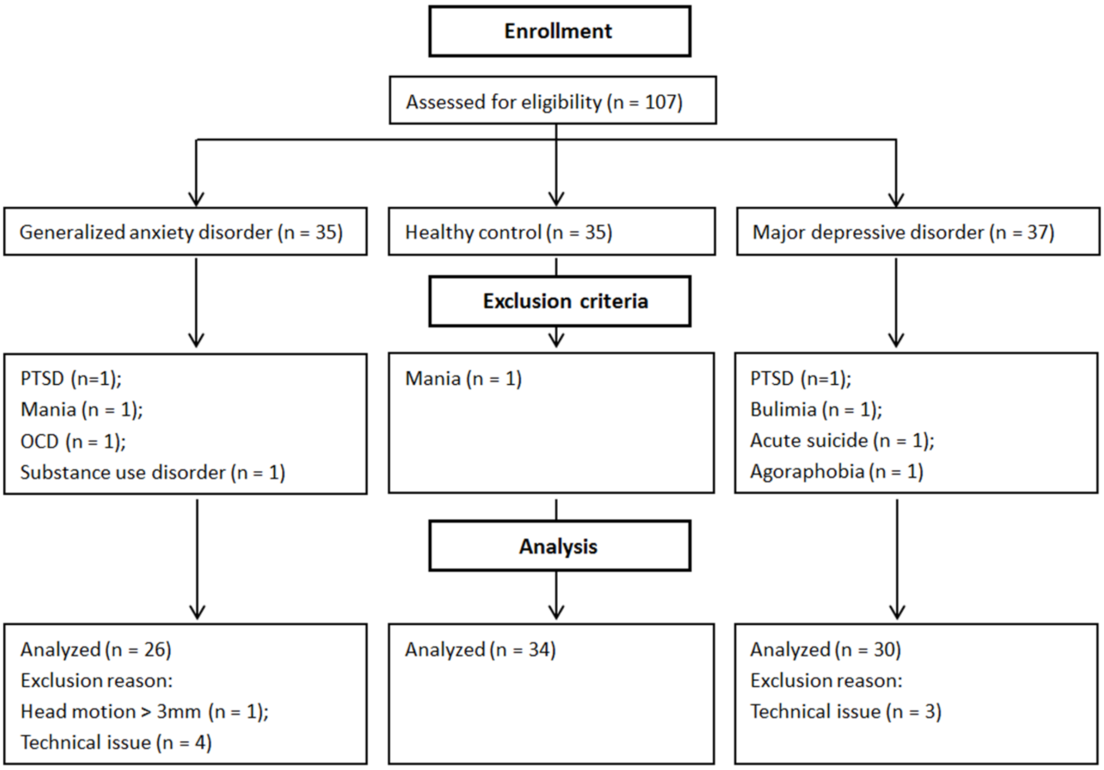
